# Random-effect based test for multinomial logistic regression: choice of the reference level and its impact on the testing

**DOI:** 10.1101/2021.04.13.21255272

**Authors:** Qianchuan He, Yang Liu, Meiling Liu, Michael C. Wu, Li Hsu

## Abstract

Random-effect score test has become an important tool for studying the association between a set of genetic variants and a disease outcome. While a number of random-effect score test approaches have been proposed in the literature, similar approaches for multinomial logistic regression have received less attention. In a recent effort to develop random-effect score test for multinomial logistic regression, we made the observation that such a test is not invariant to the choice of the reference level. This is intriguing because binary logistic regression is well-known to possess the invariance property with respect to the reference level. Here, we investigate why the multinomial logistic regression is not invariant to the reference level, and derive analytic forms to study how the choice of the reference level influences the power. Then we consider several potential procedures that are invariant to the reference level, and compare their performance through numerical studies. Our work provides valuable insights into the properties of multinomial logistic regression with respect to random-effect score test, and adds a useful tool for studying the genetic heterogeneity of complex diseases.

---

Random-effect based score test has been widely used to investigate the association between a set of genetic variants and a health outcome/trait (Wu et al., 2011; Maity et al., 2012; Sun et al., 2013). While various outcomes/traits have been considered for random-effect based score test, the multinomial outcome has received little attention until recently. Multinomial outcome analysis has important practical applications, such as the subtype analysis which concerns the association between genetic variants and multiple subtypes of a disease (Eckel-Passow et al., 2019). In multinomial analysis, one level is specified as the reference level, and the other levels are compared to this level to examine the association between the outcome and genotypes. It is generally anticipated that a statistical test should be invariant to the choice of the reference level. However, in a recent study, we made the observation that such a test in general is not invariant to the choice of the reference level (Liu et al., 2021). This is intriguing, because the logistic regression model -a model often considered as a special case of the multinomial logistic regression model -has long been observed to possess the invariance property. Moreover, the lack of invariance property for multinomial logistic regression is highly undesirable in practice, because practitioners may make potentially contradictory conclusions due to different choices of reference levels. Here, we elaborate this issue and conduct investigations to understand the fundamental cause of the problem. We first explain why the considered test is not invariant to the choice of the reference level, and then derive the analytical form of the power function when a given level is used as the reference. We next use simulations to compare several potential ways to deal with the non-invariance issue, and then provide practical guidelines at the end of the letter.

Consider a multinomial logistic regression model with *J* levels and *n* subjects. For *j* = 1, …, *J* and *i* = 1, …, *n*, let *Y*_*ji*_ = 1 if the *i*th person belongs to *j*th level, and *Y*_*ji*_ = 0 otherwise. Let *X*_*i*_ be the adjusting covariates with the first element being the intercept and *G*_*i*_ be the genotypes of *p* variants. Assume that the *J* th level is the reference level, then the model can be written as 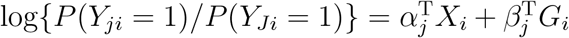, for *j* = 1, *…, J* − 1, where *α*_*j*_ and *β*_*j*_ are the regression parameters. Let *P* (*Y*_*ji*_ = 1) = *µ*_*ji*_ for *j* = 1, *…, J*. Note that 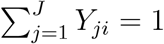 and 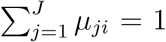. Then under *H*_0_ : *β*_1_ = *…* = *β*_(*J*−1)_ = 0, the log-likelihood can be written as

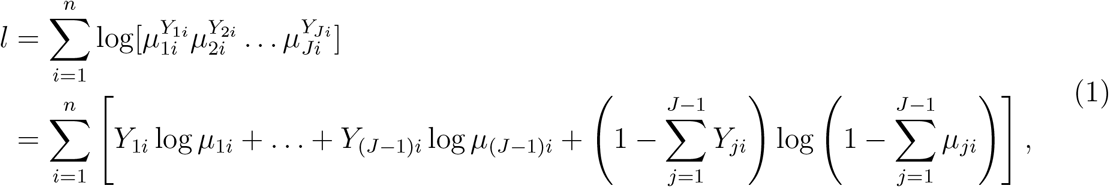

where 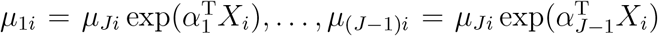. Let 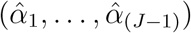 be the maximal likelihood estimator of (*α*_1_, *…, α*_(*J*−1)_) under *H*_0_. Then, the estimated *µ*_*ji*_’s are 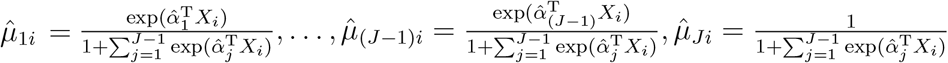. Let *G* = (*G*_1_, *…, G*_*n*_)^T^, *Y*_*j*_ = (*Y*_*j*1_, *…, Y*_*jn*_)^T^ and 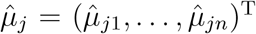. Then, the half score of the random effects for *β*_*j*_(*j* = 1, *…, J* − 1) can be derived as

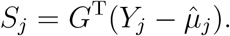

Let I_*J*−1_ = diag(1, *…*, 1)_(*J*−1)*×*(*J*−1)_, *X* = (*X*_1_, *…, X*_*n*_)^T^, 𝕏= I_*J*−1_ ⊗ *X*, 𝔾 = I_*J*−1_ ⊗ *G*, and 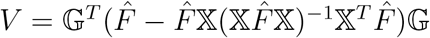, where

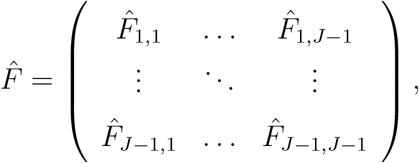

and

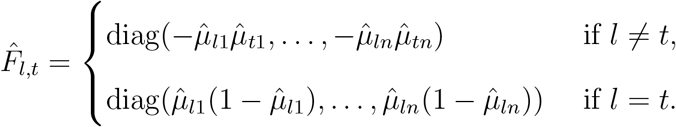

Then the score statistic 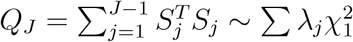, where *λ*_*j*_’s are eigenvalues of *V*. Let the p-value of this score statistic be *P*_*J*_.

Suppose that we now wish to consider a different level as the reference level. Not to lose generality, let us consider the first level as the reference level. The model can be written as 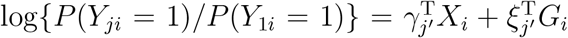, for *j* = *J*, 2, *…, J* − 1 and *j′* = 1, 2, *…, J* − 1, where *γ*_*j′*_ and *ξ*_*j′*_ are the regression parameters. Then under the null hypothesis that *ξ*_1_ = *…* = *ξ*_(*J*−1)_ = 0, the log-likelihood has a similar form as equation (1) and can be written as

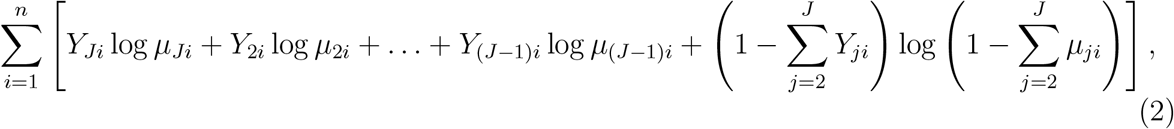

where 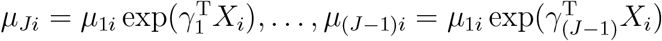. Since the likelihood in (2) is equal to that in (1), one can show that the parameter estimators

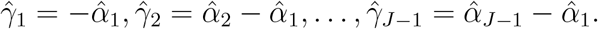

Therefore 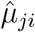 based on equation (2) is the same as that based on equation (1). Then the half score of the random effects for *ξ*_*j*_(*j* = 1, *…, J* − 1) can be written as

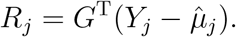

It follows that

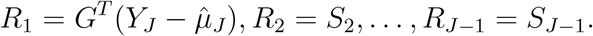

Then the score statistic 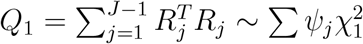, where *ψ*_*j*_’s are eigenvalues of *V*^∗^, where *V*^∗^ is the counterpart of *V*. Let the p-value of this score statistic be *P*_1_. In a similar manner, one can obtain *Q*_*j*_ and *P*_*j*_ when *j*th level is chosen as the reference level.

Recall that *S*_1_, *…, S*_*J*−1_ are the scores when *J* th level is set as the reference, while *R*_1_, *…, R*_*J*−1_ are the scores when the first level is set as the reference. The above derivation shows that there is a close relationship between *S*_1_, *…, S*_*J*−1_ and *R*_1_, *…, R*_*J*−1_. Indeed, using these results, we can further derive that

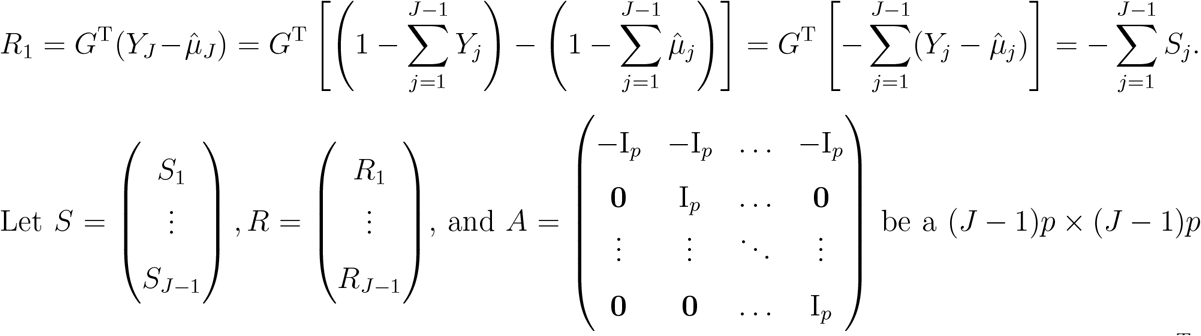

matrix, then it follows that *R* = *AS* and the covariance matrix of *R, Cov*(*R*) = *ACov*(*S*)*A*^T^. Therefore we obtain the key results that *R*^T^*R* = *S*^T^*A*^T^*AS* and *V*^∗^ = *AV A*^T^.

The above results indicate that, when a different level is chosen as the reference level, the random-effect score statistics (*Q*_*j*_ and *Q*_*j*_*′*) will have different values, and the covariance matrices for the scores will also differ. Thus, *P*_*j*_ in general is not equal to *P*_*j*_*′*. In other words, the p-values of the described statistics are not invariant to the choice of the reference level. Then an interesting question arises, that is, why does the logistic regression model, which is a special case of the multinomial logistic regression model, indeed have the invariance property? It turns out that for *J* = 2, one has *A* = −I_*p*_ and *R* = *AS* = −*S*. Then it follows that *R*^T^*R* = *S*^T^*S* and *V*^∗^ = *V*. Thus, when *J* = 2, i.e., in the case of logistic regression, the *p*-value remains the same regardless of which level is chosen as the reference level.

Since the p-value varies with the choice of reference level, we investigate how this choice influences the statistical power. For ease of presentation, let us consider *J* = 3, i.e., three levels for the outcome. Using the relationship between *R* and *S*, we have

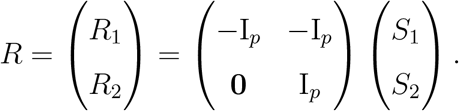

To facilitate presentation, define 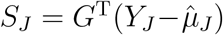, then 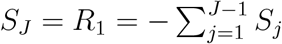. Subsequently, we have that

- the score statistic using *Y*_3_ as the reference is 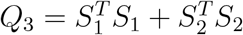;
- the score statistic using *Y*_1_ as the reference is 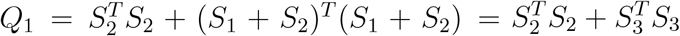;
- the score statistic using *Y*_2_ as the reference is 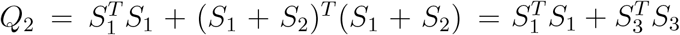.

To study the asymptotical distribution of the test statistic *Q*_*j*_ under the alternative hypothesis, let us consider a special case: *X* = 1_*n*_ ≡ (1, *…*, 1)^*T*^. Then it can be shown that 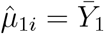 and 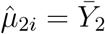. Thus

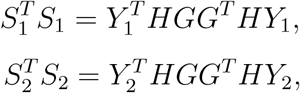

where 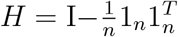. It is known that asymptotically *G*^*T*^ *HY*_1_ ∼ *N* (*G*^*T*^ *Hµ*_1_, Δ_1_ = *G*^*T*^ *H*Σ_1_*HG*), where Σ_1_ = diag(*µ*_1_(1 − *µ*_1_)). It follows that 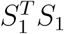 asymptotically follows a mixed noncentral chi-squared distribution:

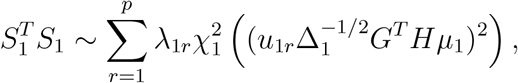

where *λ*_1*r*_’s are the eigenvalues of 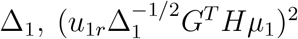 is the noncentral parameter, and *u*_1*r*_’s are the corresponding eigenvectors of Δ_1_. Similarly, let Σ_2_ = diag(*µ*_2_(1 − *µ*_2_)) and Δ_2_ = *G*^*T*^ *H*Σ_2_*HG*, then

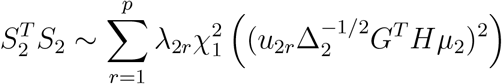

asymptotically, where *λ*_2*r*_’s are the eigenvalues of Δ_2_, and *u*_2*r*_’s are the corresponding eigenvectors of Δ_2_. To find the distribution for 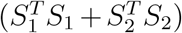, let *µ*_12_ = (*G*^*T*^ *Hµ*_1_, *G*^*T*^ *Hµ*_2_)^*T*^, Σ_12_ = *Cov*(*Y*_1_, *Y*_2_) = diag(−*µ*_1_*µ*_2_), and

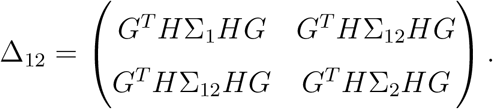

Then one can derive that

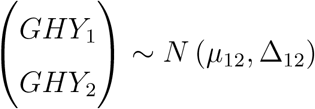

asymptotically. Therefore, 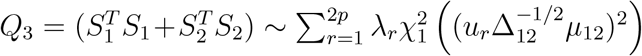 asymptotically, where *λ*_*r*_’s are eigenvalues of Δ_12_, and *u*_*r*_’s are the corresponding eigenvectors of Δ_12_. In a similar manner, we can derive the asymptotical distributions of *Q*_1_ and *Q*_2_, respectively.

Recall that the power function is Ψ_*j*_(*Q*_*j*_ *≥ c*_*j*_), where Ψ_*j*_ is the cumulative distribution function of *Q*_*j*_ under *H*_1_, and *c*_*j*_ is the critical value determined by the distribution of *Q*_*j*_ under *H*_0_. It is tempting to directly compare *Q*_*j*_’s power functions using the above derived asymptotical distributions, but it is challenging to do so. This is because when the reference level is replaced, the *Q*_*j*_’s asymptotical distributions under both the null and the alternative hypotheses will change, making it extremely difficult to compare the power across difference reference levels. On the other hand, when the *J* subtypes have similar proportions among the *n* subjects and there is no adjusting covariate, one can show that the asymptotical distributions of the *Q*_*j*_’s are approximately equal to each other under the null hypothesis. Then it follows that the larger the statistic *Q*_*j*_ is, the more likely one will reject the *H*_0_. Recall that 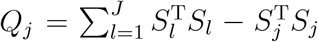. This suggests that, to maximize the power, the level with the smallest 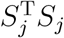 should be chosen as the reference. Our simulation studies confirmed this derivation (see Supplementary Material). The size of 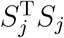 has practical interpretations. Recall that *S*_*j*_ is the inner product between 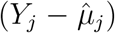 and *G*. Thus, 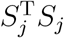 can be roughly seen as the correlation between *Y*_*j*_ and genotype. This suggests the level that has the weakest correlation with the genotype should be chosen as the reference level, which well matches intuitions.

The above analysis provides theoretical insights into the power of the random effects score test. However, in practice, it is generally unknown which 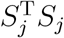 is the smallest among the *J* levels. This can be seen from the following. Taking *S*_1_ as an example, we have

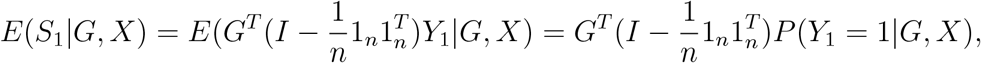

where *P* (*Y*_1_ = 1|*G, X*) is a *n*-length vector with each element being

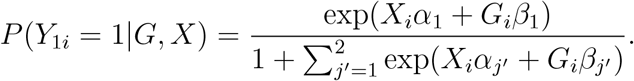

Clearly, *E*(*S*_1_|*G, X*) is a quantity related to *G, X, α*_1_, *α*_2_, *β*_1_, *β*_2_. Since *α*_1_, *α*_2_, *β*_1_, *β*_2_ are unknown parameters, it is difficult to evaluate the size of 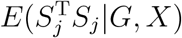 accordingly. Hence, practical data analysis will need statistical tests that are invariant to the choice of the reference level. In the following, we consider three procedures to tackle this issue, and compare the performance of the three methods through simulation studies.

## I. A Bonferroni procedure

We use each of the *J* levels as the reference level, and based on *Q*_*j*_’s, obtain the corresponding p-values *P*_1_, *…, P*_*J*_. Then, use 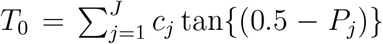 as the final p-value. The multiplication of *J* is a Bonferroni correction to ensure that correct type I error is maintained.

## II. A Cauchy procedure

We propose to adapt to a Cauchy procedure (Liu and Xie, 2020) to combine the *J* p-values, *P*_1_, *…, P*_*J*_. Specifically, let 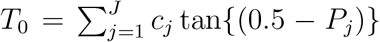, where 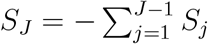 and *c*_*j*_ is the pre-specified weight to accommodate prior knowledge on *j*th level. When there is no prior knowledge on the *J* levels, all *c*_*j*_ = 1*/J*. Then under the null hypothesis, the p-value of *T*_0_ can be approximated by (1*/*2 − (arctan *T*_0_)*/π*) based on the Cauchy distribution.

## III. An integrative procedure

Consider a statistic *L* = (*WDS*)^*T*^ (*WDS*), where *W* = diag(*w*_1_, *…, w*_*J*_) ⊗ I_*p*_, *D* = (I_*J*−1_, −**1**_*J*−1_)^*T*^ ⊗ I_*p*_, and *w*_*j*_ is a pre-specified weight for *j*th level. When *w*_*j*_’s are all equal, this statistic reduces to a statistic in Liu et al. (2021). Using the relationship that 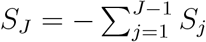, we can show that 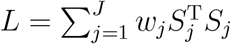, which is invariant to the choice of the reference level. Alternatively, *L* can be written as 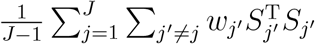, where 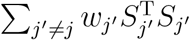 is a weighted version of *Q*_*j*_. Thus, *L* can be seen as an integrative statistic that consists of all the *Q*_*j*_’s. *L* asymptotically follows 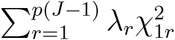, where *λ*_*r*_ are the eigen values of *WDV D*^T^*W* ^T^, and and 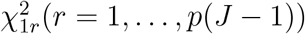 are independent 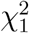 random variables.

We conducted simulation studies to examine the type I errors of these procedures. We considered three levels for the response variable, and generated an adjusting covariate from *N* (0, 1). The regression coefficients for the intercept and the adjusting covariate were set as *γ*_1_ = (0.3, 1.2)^T^ and *γ*_2_ = (0.3, 0.9)^T^. Next we simulated a *p*-vector of mutations with each element generated from a Bernoulli(0.05). To examine the type I error, we set *ξ*_*j*_ = **0** for *j* = 1, 2 and considered *n ∈* {300, 500, 1000} for *p* = 10, 15. We evaluated the type I error at significance level *α* = 10^−3^. A total of 10^6^ simulated datasets were generated for each setting. As shown in Table 1, all considered procedures are able to control the type I error. Next, we examined the power of these procedures. We considered two scenarios:

**Table 1:**
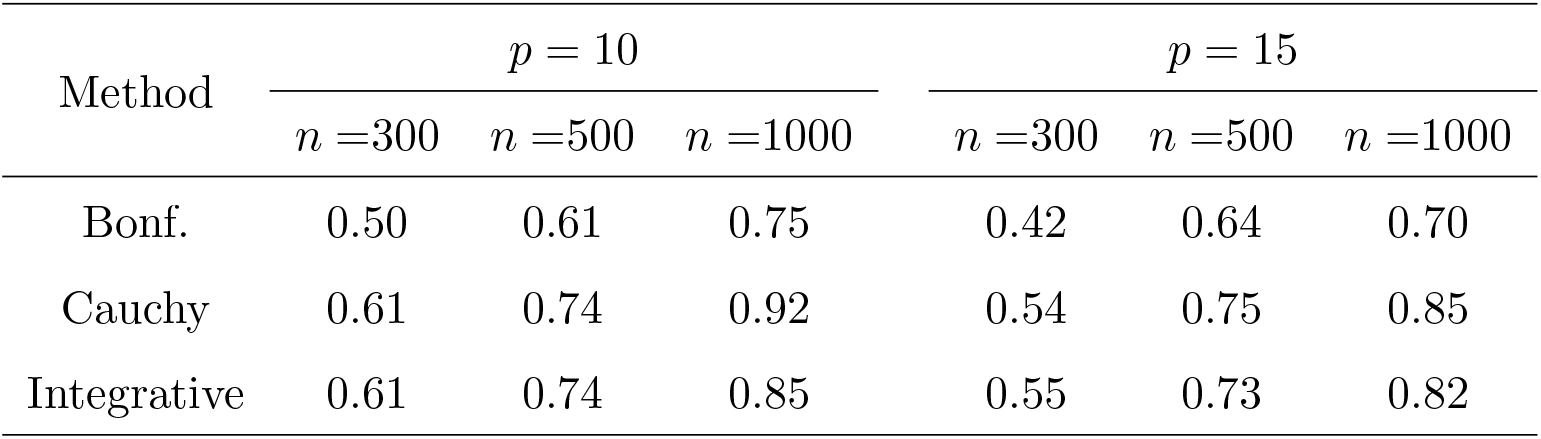
Empirical type I error (*×*10^−3^)

1. 60% of *ξ*_*js*_’s were generated from Uniform(0.3, 1.5), and 40% of *ξ*_*js*_’s were generated from Uniform(−1.5, −0.3).
2. 60% of *ξ*_*js*_’s were generated from *N* (0, 1.4^2^), and 40% of *ξ*_*js*_’s were set to 0.

*ξ*_*js*_’s were fixed over all replicates. Each scenario was replicated 10^4^ times. The power for scenarios I and II is summarized in Tables 2 and 3. The Bonferroni procedure has the lowest power, due to its conservativeness in controlling type I error. The integrative procedure properly accounts for the correlations among the *J* statistics, and tends to have the best performance among the considered procedures. Thus, we recommend the integrative procedure for practical data analysis.

**Table 2:** Power for scenario I

| Method | $p = 10$ | | $p = 15$ | |
| --- | --- | --- | --- | --- |
| | $n = 250$ | $n = 300$ | $n = 250$ | $n = 300$ |
| Bonf. | 0.45 | 0.64 | 0.86 | 0.96 |
| Cauchy | 0.49 | 0.68 | 0.89 | 0.97 |
| Integrative | 0.55 | 0.74 | 0.90 | 0.98 |

**Table 3:** Power for scenario II

| Method | $p = 10$ | | $p = 15$ | |
| --- | --- | --- | --- | --- |
| | $n = 250$ | $n = 300$ | $n = 250$ | $n = 300$ |
| Bonf. | 0.55 | 0.76 | 0.86 | 0.96 |
| Cauchy | 0.60 | 0.80 | 0.89 | 0.97 |
| Integrative | 0.66 | 0.84 | 0.93 | 0.98 |

In summary, we have shown that the random-effect score test for multinomial logistic regression is not invariant to the choice of the reference level. Our results provide analytical explanation to this issue, and simulation studies confirmed that the choice of the reference level influences the statistical power. We considered several procedures that can yield p-values (or statistics) that are not dependent upon the reference level, and the integrative procedure appears to have a more favorable performance. Overall, our study provides new insights into the random-effect score test for multinomial logistic regression, and will aid in the ongoing study of genetic heterogeneity for complex diseases.

## Data Availability

Data used in this manuscript are simulated.

## Notes

### Competing Interest Statement

The authors have declared no competing interest.

### Funding Statement

This work was supported by NIH grant R01CA223498 and R01CA189532.

### Author Declarations

All data are simulated. No real data are involved in this manuscript.

