## Supplementary Figure S1 for "Random-effect based test for multinomial logistic regression: choice of the reference level and its impact on the testing"

### Relationship between the size of $\|S_j\|$ and the power

We conducted simulation studies to examine the relationship between the size of $\|S_j\|$ and the power. Here, the $\|S_j\|$is defined as $(S_j^T S_j)^{0.5}$. We generated an adjusting covariate from Bernoulli(0.5). The regression coefficients for the intercept and the adjusting covariate were set as $\gamma_1 = (0.3, -0.6)^T$ and $\gamma_2 = (0.2, -0.4)^T$. We considered the following random effects in the model: 50% of $\xi_{1s}$'s were generated from $N(0.3, 1)$, and 50% of $\xi_{1s}$'s were generated from $N(-0.3, 1)$; 50% of $\xi_{2s}$'s were generated from $N(0.2, 1)$, and 50% of $\xi_{2s}$'s were generated from $N(-0.2, 1)$. These $\xi_{js}$'s were fixed over all replicates. We considered $n = 300$, $p = 10$, and 500 replicates.

The boxplot of $\{S_j^T S_j, j = 1, 2, 3\}$ is presented in Figure S1. Under the above simulation set up, we observed that $S_2^T S_2$ tends to have the highest value, followed by $S_1^T S_1$ and $S_3^T S_3$. Correspondingly, the power of $Q_1, Q_2$ and $Q_3$ was observed to be 0.740, 0.574 and 0.892, respectively. These results show that, when the level with the lowest $S_j^T S_j$ is chosen as the reference level, one tends to have the highest power for the association test.

Supplementary Figure S1: Boxplot of $S_1^T S_1, S_2^T S_2$, and $S_3^T S_3$

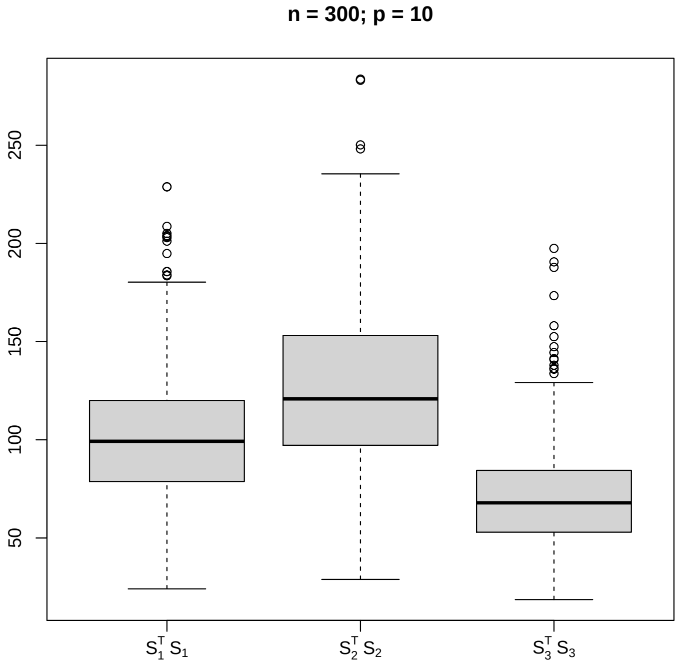
